# Estimating the lagged effect of price discounting: a time-series study using transaction data of sugar sweetened beverages

**DOI:** 10.1101/2021.09.26.21264133

**Authors:** Hiroshi Mamiya, Alexandra M. Schmidt, Erica E. M. Moodie, David L. Buckeridge

**Author notes:** **Contact information:** Hiroshi Mamiya, 11-4119 Ave Madison, Montreal, QC, H4B 2T8, Canada, Fax: Currently unavailable. **Author contributions:** The study was conceived and designed by Mamiya and was reviewed and approved by the other authors. Schmidt and Moodie provided inputs on the statistical analysis and interpretation of the results. Buckeridge provided the data and computational resources. Data analysis and drafting of manuscript was led by Mamiya. All authors reviewed, provided critical comments to the manuscript, and approved the final version of the manuscript for submission. **Ethics of human subject participation:** This study used secondary data that are aggregated store-level measurements of consumer purchasing, rather than individual consumer level data. The study therefore did not require a separate written or verbal consent from human subjects.

## Abstract

**Background:** Price discount is an unregulated obesogenic environmental risk factor for the purchasing of unhealthy food, including Sugar Sweetened Beverages (SSB). Sales of price discounted food items are known to increase during the period of discounting. However, the presence and extent of the lagged effect of discounting, a sustained level of sales after discounting ends, is previously unaccounted for. We investigated the presence of the lagged effect of discounting on the sales of five SSB categories, which are soda, fruits juice, sport and energy drink, sweetened coffee and tea, and sweetened drinkable yogurt.

**Methods:** We fitted a distributed lag model to weekly volume-standardized sales and percent discounting generated by a supermarket in Montreal, Canada between 2008 and 2013.

**Results:** While the sales of SSB increased during the period of discounting, there was no evidence of a prominent lagged effect of discounting in four of the five SSB; the exception was sports and energy drinks, where a posterior mean of 28,459 servings (95% credible interval: 2,661 to 67,253) of excess sales can be attributed to the *lagged* effect in the target store during the study period.

**Conclusions:** Our results indicate that previous studies may have underestimated the effect of price discounting for some food categories.

**Key messages:** *What is already known:* Temporary price discounting is an important component of obesogenic food environment, as it has been shown to increase the sales of discretionary food items during the period of discounting.

*Novelty of findings:* Even after a period of price discounting has ended, the sales of sports and energy drinks remain at a higher level relative to the sales before discounting.

*Research and public health implications:* Previous research focusing on the immediate effect (i.e., same time period) of price discounting may have systematically underestimated the impact of price discounting for some food categories. The findings and analytical method in this study promote improved validity of future food environment research targeting the impact of discounting and other types of food promotions on the sales of energy-dense and nutrition-poor food items.

## Introduction

Sugar Sweetened Beverages (SSB) represent the largest source of dietary sugar in many nations ^1^ and are epidemiologically linked to obesity, overweight and nutrition-related chronic diseases ^2^. Price discounting, the temporary reduction of price per unit food, is one of the most important marketing tactics used by food retailers and manufacturers to increase sales ^3,4^. Prevalence of price discounting is often reported to be disproportionately higher among highly processed ‘junk’ food including SSB ^5^, and people’s purchasing of SSB appears to be particularity susceptible to price discounting –more so than solid (non-beverage) food ^6,7^. Price discounting may lead to the overconsumption of the promoted food items ^3,8^, thus being a retail (in-store) environmental risk factor for food diets inconsistent with nutrition guidelines.

From an intervention perspective, price discounting is a highly unregulated and neglected environmental risk factor for unhealthy eating ^9^, as the only regulatory initiative to date is the UK and Scotland government’s proposal for the mandatory restriction of volume-based discounting (e.g. reduced price for multi-buy) and display promotion of price-discounted food items ^8^. Given the lack of interventions and natural experiment to ban price discounting, evidence from observational studies characterizing the impact of discounting on population nutrition may provide motivative knowledge for governmental actions in other settings.

Several pioneering studies in public health nutrition found an association between discounting and increased sales of the promoted food items, mainly based on cross-sectional analyses that pooled purchasing and discounting records during the entire study period^5,10,11^. The findings are confirmed by the results from longitudinal studies controlling for time-varying confounders (e.g., season and other forms of time-varying marketing activities) including our previous work ^3,12,13^. While the increase of sales during the period of discounting is consistently observed, time-lagged effect of discounting, or the association of discounting at current time with sales in the post-discounting time periods, has not received research attention.

A lagged effect of marketing exposure, including price discounting, can occur due to repeated purchasing of items where initial sales were triggered by marketing activities. Although the lagged effect of price discounting were investigated and confirmed by marketing researchers for some food categories ^3,14,15^, these findings do not readily apply to food groups of public health interest e.g., beverages may not be separated into diet (without artificially added sugar) and their non-diet (SSB) counterparts and often focus on sales for a small number of top-selling brands within a food groups of interest ^15^. One study conducted by a marketing firm for Public Health England suggest the potential lack of such effect ^16^; however, no longitudinal studies in public health nutrition specifically targeted the identification of lagged effects of price discounting (and cross-sectional studies are, by design, unable to estimate the temporal lag of an exposure effect). Lagged effect therefore remains as a potentially important source of bias in the association of price discounting (and other promotional activities, such as display and flyer promotions) with sales, potentially leading to an underestimated association.

The objective of this study is to conduct a time-series analysis to assess the presence and magnitude of a lagged effect of discounting for five SSB categories based on weekly time-series of retail transaction data in a large supermarket in Montreal, Canada. The SSB categories of interest are 1) carbonated soft drinks (hereafter termed soda), 2) fruits drinks (less than 100% fruit beverages), 3) sports and energy drinks, 4) sugar-sweetened coffees and teas, and 5) sugar-sweetened drinkable, as opposed to spoonable, yogurt. These are non-alcoholic beverages containing artificially added sugars and not containing artificial sweeteners, thus excluding diets products. This is to our knowledge the first study to provide insights about the lagged effect of within-store obesogenic marketing activities.

## Methods

### Study design

This is a retrospective time-series study investigating the association between weekly discounting and sales of five SSB categories in a single supermarket located in Metropolitan Montreal, Canada. The study time period is between January 2008 and December 2013, thus consisting of 311 weeks, or 6 years. The unit of analysis is weekly sales transactions for each beverage category. Note that this is not a longitudinal data analysis that uses measurements from multiple stores as seen in our previous studies ^12,13^, i.e. these are not panel data. Rather, we perform a time-series (i.e., single store) analysis, which allows us to explore time-lagged effects while accounting for temporal correlation of sales.

### Transaction data

The transaction records were generated by a large supermarket owned by a major Canadian retail chain (the identity of the chain is anonymized) and were purchased from a marketing firm, Nielsen ^17^. The store neighborhood, as defined by Forward Sortation Area (first 3 digits of Canadian postal codes) in which the store was located, had median income of 68 627 Canadian dollars (median of income of all areas in Metropolitan Montreal was 71 906 [IQR: 56 580 – 83 840], according to the 2011 Canadian National Household Survey) ^18^.

The data consist of weekly sales quantity of individual beverage items, as uniquely defined by the Universal Product Code and item name, weekly price of sold items in Canadian cents, flyer promotion and retail display promotion (described below). We classified these items into the five non-alcoholic SSB categories based on product name of each beverage item and corresponding food category assigned by Nielsen. For example, soda items were categorised by the company as “carbonated soft drink”, but we manually excluded diet soda i.e., items with artificial sweeteners based on terms such as “diet”, “zero”, “non-sugar”.

### Outcome

The weekly sales quantities of each beverage item were standardized to the Food and Drug Administration’s single serving size of 240ml for beverage (approximately 1 cup). The outcome variable is the aggregated sum of sales from items in each category in each week, where the category-specific average number of distinct items over the entire 6 year period in our store was 109 (soda), 152 (fruits drinks), 36 (sports and energy drinks), 22 (coffees and teas), and 29 (drinkable yogurts). The category -specific sales were natural log transformed to reduce skewness. We did not analyse the disaggregated, individual item-level association between sales and discounting, since such an analysis required us to account for across-item dependency of sales. Since the change of category-level sales is the primary interest to population nutrition over the sales of individual food item or brand, our unit of analysis for both exposure, outcome and confounders was defined at the level of beverage category.

### Exposure

The exposure variable is the binary indicator of category-specific discounting at each week. It was calculated as the weighted average of weekly price discounting of items in each category, with weights representing each item’s market share (proportion of serving-standardized sales) within the category it belongs to. Price discounting of an individual food items was calculated as percent decrease of the serving-standardized price sold (net price) from the baseline (i.e., non-promoted) price ^12,19^. Detailed calculation of serving-standardized discounting for each item and subsequent aggregation to category is provided in Appendix S1 in Supplementary Information File.

### Statistical analysis: regression variables to capture lagged association of price discounting and SSB sales

A lagged association between time-varying outcome (log-transformed sales quantity) and exposure (discounting) is commonly captured by a distributed lag model, which is a regression model that contains multiple time-lagged values of an exposure. Regression coefficients for these time-lagged variables have functional constraints (i.e., the value of the coefficients is constrained to change smoothly over lag) as frequently seen in environmental time-series epidemiology and econometrics^20,21^. One such constraint is the Koyck lag decay ^22^, which captures the monotonic decay of the effect of an exposure over time by two regression coefficients: *β* as the immediate effect (at lag zero) and *λ* as the lag coefficient that quantifies the decaying rate. The functional form of the Koyck decay is represented by the polynomial of form:

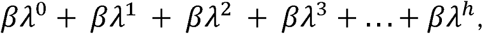

where *h* indicates lag, and *β λ*^0^ = *β* is the immediate effect. An estimated value of the lag coefficient *λ* closer to 0 indicates the absence of lag, while its value closer to 1 indicates a stronger lag effect. The visual interpretation of the lagged effect represented by this polynomial function is provided in Supplementary Figure 1. We pre-specified the range of the estimated value of *λ* to be 0 < *λ* < 1 so that the effect of discounting monotonically decays towards zero over lag, capturing a diminishing effect.

**Figure 1:**
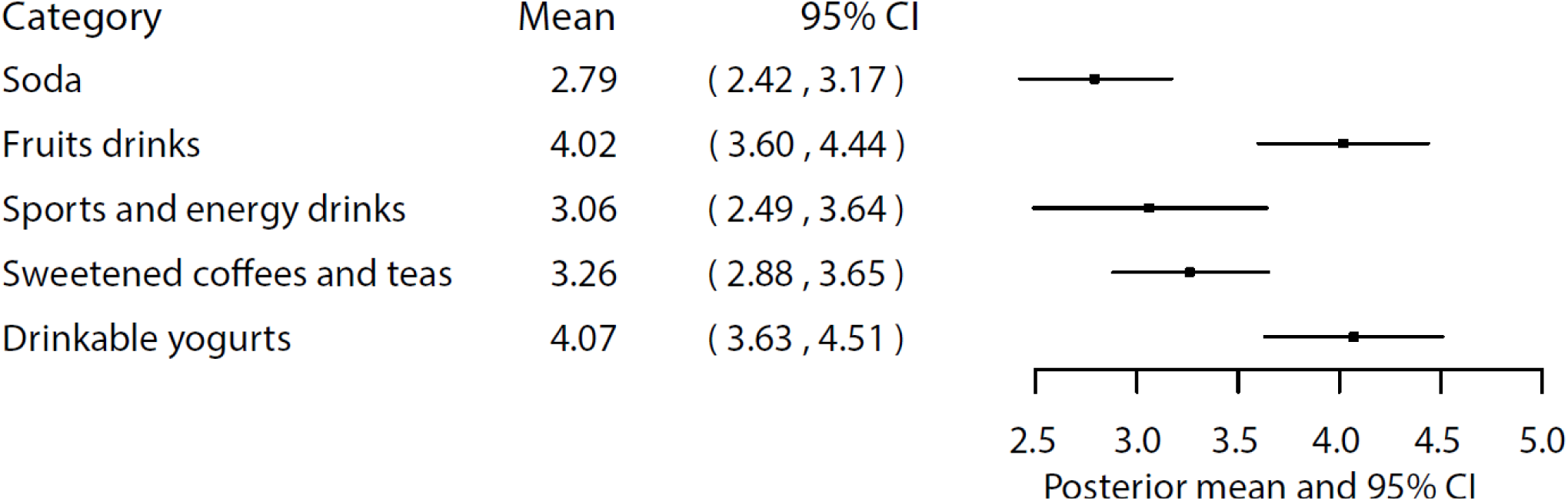
The estimated immediate effect for price discounting on the sales of five sugar-sweetened beverage categories. The value represents the percent increase of non-log sales upon one percent discounting for each SSB category, as calculated by multiplying the posterior summary of by 100.

### Statistical analysis: time-series regression model to incorporate Koyck lag model

The Koyck lag variables were added to a linear time-series regression, dynamic linear model^23,24^. The details of the model, including the intercept and the lag coefficients, are provided in Appendix S3. Covariates are weekly varying variables that are likely to temporarily correlate with price discounting and sales. These include non-discounting promotion: display promotion and flyers, which often co-occur with price discounting (although not always) and associated with higher sales^3^. Display promotion is temporarily placement of items into prominent location of stores such as store front. As in price discounting, we calculated the value of these variables at the level of SSB category by aggregating binary promotion status across items. Specifically, display promotion is coded as 1 if an item was temporarily placed at any one of prominent retail locations from the original shelf space, such as the end of aisle, entrance to store, or by the cashier. Flyer promotion was coded as 1 if an item was listed in flyer, and 0 otherwise. These item-level binary variables were aggregated to the category-level proportion as the weighted proportion of items promoted in each category at a given week, where the weights represented an item’s serving-standardized market share, as in the discounting variable.

We also added covariates for seasonal trends that were defined as the sine- and cosine-transformed harmonic wave of a week variable as detailed in Appendix S3. Additionally, an indicator variable for week containing national and provincial statutory holidays were added. Other variables included in the models were regular (baseline) price of each beverage categories, mean daytime temperature in each week, and the lagged value of sales itself (autoregressive of order 1). Note that autoregressive terms included in this way in a time series analysis are not equivalent to those that might be used in panel data in the context of longitudinal data analysis (i.e., linear mixed model)^12,13^.

We fitted a separate model for each of the five food categories independently under the Bayesian framework. We therefore specified prior distributions for regression parameters (Appendix S3). Interpretation of regression coefficients is based on point estimates (posterior mean or median) and uncertainty (95% Credible Interval [CI]) as summarized from the posterior distribution of the parameters simulated by Markov Chain Monte Carlo methods. Model selection, specifically selecting covariates was guided by the value of the Watanabe-Akaike Information Criterion (WAIC) indicator of model fit^25^. A lower WAIC value indicates a better-fitting model. Codes are publicly available: https://github.com/hiroshimamiya/promotionLag/blob/main/discountLag_KoyckTransfer.stan.

As a sensitivity analysis, we tested an alternative shape of promotion decay by changing the constraint of the lag parameter *λ* from 0 < *λ* < 1 to −1 < *λ* < 0. The latter specification implies that, rather than assuming monotonic decay seen in Supplementary Figures S2 a and b, we allowed the model to capture a so-called ‘post-promotion dip’ (Supplementary Figures S3), a sharp reduction of sales below pre-discounting period immediately after discounting ^3^. Thorough theoretical explanations for the post-promotion dip are provided elsewhere ^3,26,27^.

## Results

### Descriptive analysis

The median sales quantity of the SSB categories across 311 weeks in the target store varied widely across the SSB categories, with soda and fruit drinks being the largest source of SSB sales (Table 1). These two categories, however, exhibited a decreasing trend during the study period (Supplementary Figure S4 a and b), consistent with the trend estimated by the 2004 and 2015 Canadian national nutrition survey ^28^. The sales of sports and energy drinks exhibited strong seasonal (cyclic) patterns (Supplementary Figure S4 c), but the sales in this store did not appear to show the increasing trend of sales that has been observed in Metropolitan Montreal, Canada and worldwide in the same time period and reported elsewhere ^13,28,29^. Discounting of fruit drinks and sweetened coffees and teas rose steadily over time (Supplementary Figure S5 b and d). Average percent discounting over the study period was highest for soda and lowest for yogurt (Table 2).

**Table 1.** Summary of weekly sales quantities of SSBs in the target store between 2008 and 2013, in non-log scale serving quantity.

| SSB category | Mean | Median | IQR |
| --- | --- | --- | --- |
| Soda | 20,330.7 | 19,062.5 | 14,057.4, 25,351.9 |
| Fruits drinks | 14,237.9 | 14,303.5 | 10,887.3, 17,092.1 |
| Energy and sports drinks | 1,400.4 | 1,178.4 | 869.5, 1,763.2 |
| Sweetened coffees and teas | 1,514.5 | 1,397.0 | 1062, 1,825.3 |
| Sweetened drinkable yogurts | 1,368.8 | 1,226.7 | 991.4, 1,594.5 |

Abbreviations: IQR, Interquartile Range

**Table 2.** Summary of weekly percent discounting per serving of SSBs in the target store between 2008 and 2013.

| SSB category | Mean | Median | IQR |
| --- | --- | --- | --- |
| Soda | 18.0 | 17.3 | 12.6, 24.3 |
| Fruits drinks | 11.8 | 9.8 | 5.5, 16.3 |
| Energy and sports drinks | 8.4 | 8.2 | 1.0, 12.5 |
| Sweetened coffees and teas | 15.5 | 13.8 | 7.6, 21 |
| Sweetened drinkable yogurts | 6.3 | 4.4 | 2.0, 9.5 |

Abbreviations: IQR, Interquartile Range

### Time-series regressions

The summary of the time-varying intercept for each of the SSB categories shows its temporal path capturing the local fluctuations and overall declining trends of the SSB sales, as seen in Supplementary Figure S6 a-e. Inspection of autocorrelation plots of residuals suggests little serial correlation (Supplementary Figure S7 a-e).

### Immediate effect of discounting

The estimated coefficient *β* indicating the change of sales during the time of discounting is shown in Figure 1. The values represent the change in natural log-transformed serving-standardized sales quantity by one percent increase of price discounting during the period of discounting, which is equivalent to the *percent* change of non-log sales when multiplied by 100. This immediate effect was highest for drinkable sweetened yogurt and lowest for soda (Figure 1). In other words, sweetened drinkable yogurt category is subject to the highest “deal-proneness” among the five SSB categories in this store. Soda beverages showed the weakest immediate effects.

### Lagged effect of discounting

The extent of lagged effect (the coefficient λ) for each food category is provided in Figure 2. The estimated value of *λ* is close to zero for all SSB categories but somewhat larger for sports and energy drinks. The visual interpretation of the lagged effects in the form of the above mentioned Koyck polynomial function for each SSB category (Figure 3) indicate that the percent increase of sales relative to baseline (pre-discounting period) immediately drop to almost zero at lag 1. These shapes suggest a diminishing effect immediately after the period of discounting (i.e., lag 0), except for a weak and therefore short-term lagged effect for sports and energy drinks.

**Figure 2:**
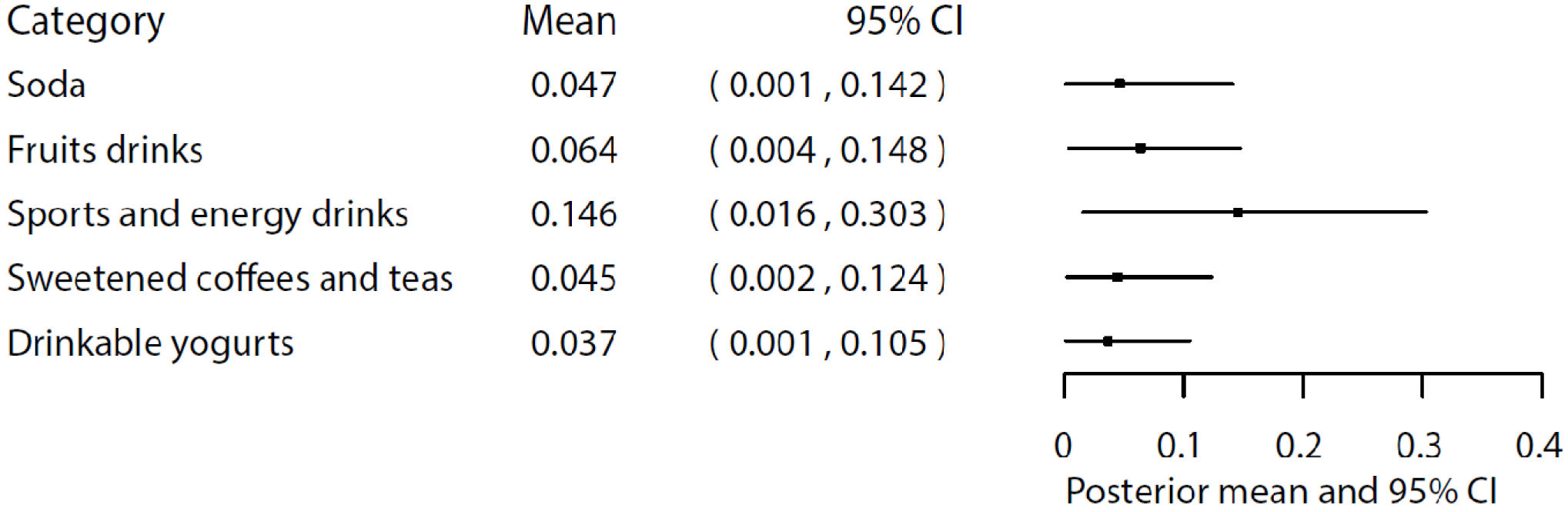
Posterior summary of the estimated lag coefficient,, for price discounting on the sales of five sugar-sweetened beverage categories

**Figure 3.**
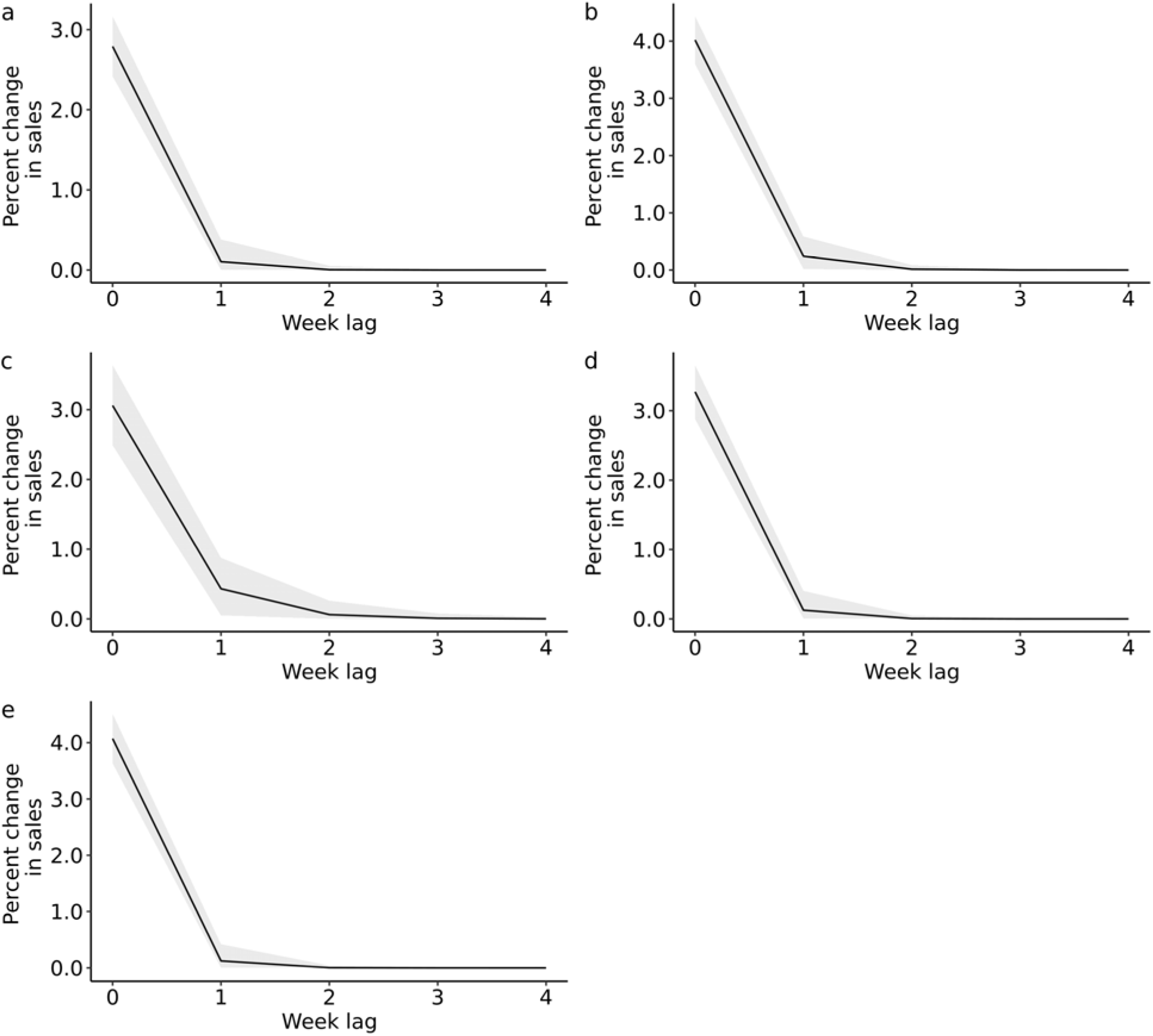
Impulse response function showing the lagged effect of price discounting on the sales of a) soda, b) fruit drinks, c) sports and energy drinks, d) coffees and teas, and e) drinkable yogurts sports and energy drinks, with 95% Credible Interval indicated by the gray polygon. The value at *x* = 0 represents the immediate effect represented by the posterior median of *β*, the percent change of sales during the period of one percent discounting.

We also quantified the absolute total excess sales of each SSB categories attributable to the lagged effect alone over the 6 years of observation of discounting in our store (Supplementary Table S2). This is the posterior distribution of the difference between exponentiated fitted sales generated by the model containing the lag parameter λ and the exponentiated fitted sales generated by the model with the immediate discounting effect alone (i.e., setting λ= 0). For sports and energy drinks, these excess sales due to lagged effects of discounting are summarized by a posterior mean of 28,459 (median = 26,345, with 95% CI 2,661 to 67,253) servings, which is approximately 21% of the sales quantities attributable to the total (immediate plus lagged effect of price discounting combined, posterior mean = 131,606, median = 130,446, 95%CI = 96,155 to 173,625).

A sensitivity analysis to inspect the presence of the post-promotion dip effect using alternative constraints of the lag coefficient (−1< λ< 0) showed inferior model fit to the original specification (0< λ < 1) which modelled a monotonic decay of discounting effect.

## Discussion

We investigated time-lagged effect of price discounting for five SSB categories for a supermarket located in Metropolitan Montreal, Canada. The results indicate that the association between discounting and sales of sports and energy drinks persisted even after discounting ended. To the best of our knowledge, the extant public health research estimating the association of price discounting and sales has evaluated only the immediate effect, thus potentially leading to a biased (underestimated) association of price discounting with sales of some food categories.

There is an increasing number of studies investigating within-store food promotions as a modifiable obesogenic environmental drivers of (un)healthy food selection and nutrition disparities ^11^, and price discounting is likely to have the most influential impact on food purchasing ^4,15^. Similar to the exposure to environmental hazard (e.g., pollution, heat wave), lagged effect of marketing exposure should be considered to be one of potential sources of bias, as seen in this study and literature in marketing science ^3^, as well as recent research investigating the impact of media advertising on population nutrition ^30^. While it is reassuring that the lagged effect is absent for the SSB categories such as soda in the store investigated, the presence of such effect for the sales of sports and energy drinks implies potential underestimation of the influence of discounting in previous studies targeting these beverages, including ours ^13^.

The lagged effect on the SSB category of sports and energy drinks may have occurred due to repeated trials induced by discounting among peoples who are previously unexposed to the consumption of these rapidly expanding SSB beverages. Sales and consumption of these beverages, in particular energy drinks, exhibited a steady and global growth during the study period ^13^, mainly propelled by aggressive and ubiquitous marketing activities within and outside retail settings, including sponsoring of sports and youth events ^31–33^. While the percent increase of sales due to the lagged effect appears modest relative to the immediate effect, the absolute quantity of sports and energy drinks attributable to the lag effect is concerning. Aside from their sugar contents, a single serving of energy drinks often reaches the recommended daily dose of caffeine intake among youth ^34^ and associated with caffeine-related acute health outcomes including neurological, psychological and often fatal cardiovascular events ^33,35^.

Possible reasons for the absence of discounting carryover effect in the other SSB categories include rationale planning of shopping activities i.e., not buying items until next promotions ^3^. This forward-looking planning may be relevant for categories that are discounted heavily, namely soda, fruit drinks and coffees and teas as seen in the descriptive analysis. It is also possible that the lagged effect is masked by the aggregated measure of sales and discounting by SSB categories in this study. In other words, individual food items within categories may exhibit a lagged effect, but the increased sales due to such effects maybe an expense of reduced sales on competing items within the same category – often termed as “cannibalization” due to people’s switching of food items within a category ^3^. Thus, the overall category sales might not have increased at post-discounting period. This explanation also applies to the results of the sensitivity analysis: the lack of post-promotion sales dip frequently observed in the disaggregated brand-level analysis ^26,27^.

Our findings should be interpreted with several limitations in mind. First, while one of the key contributions of this study is to introduce an exposure lag model applicable to other populations, the data in this study are not very recent (2008-2013). As well, our findings are based on shopping patterns in a single supersmarket. Population-level influence of discounting across varying socio-economic status at the shopper- or store neighborhood-level needs to be estimated based on a regionally representative and more recent sample of stores or people. This would require panel data, which in turn would bring significant increases in the computational and analytical complexity, requiring hierarchical analyses of lagged models with spatial correlation across geographical locations of stores. As in any observational study, we note the potential for unmeasured confounder of price discounting, such as media advertising. Finally, it is possible that potential switching of SSB purchasing in nearby stores led to measurement error in our store, although the proportion of individuals prone to switch shopping venues appears to be relatively small (10-15%) and this pattern of store-substitution more typically occurs for high-cost items such as coffee and beer.^36,37^

Future research should investigate the lagged effect of other forms of sales promotions, including couponing, volume discount, display and flyer promotions, which independently and jointly influence selections of energy-dense and nutritionally poor food items.^3,11^

## Conclusion

Overall, our results provide insights into the lagged effect of price discounting on unhealthy beverage purchasing that should be further investigated by other observational studies, as such effect may represent a previously overlooked source of bias in the association of sales and within-store food marketing activities, which is recognized as a potentially important but largely unregulated component of obesogenic food environment. We remark that, while the analytical approach provided in this study is a flexible form of distributed lag model (no need to specify the lag length *a priori*), there are alternative and readily implementable regression models to capture lagged effects, many of which are distributed as software library (typically from the frequentist statistical point of view) and commonly appear in other domains of public health with extensive documentation ^38–40^.

## Data Availability

Scanner transaction data form retail food outlets are collected in many nations by the Nielsen company. The data are available through commercial agreement with the company or through affiliated academic institutions that maintain access to these data for research use.

