## Supplementary File for "Estimating the lagged effect of price discounting: a time-series study using transaction data of sugar sweetened beverages"

### Appendix S1. Detailed Description of Calculation of Category-Level Discounting

As in previous studies, we used a conventional marketing approach to calculate the category-level discounting of Sugar Sweetened Beverages (SSB) for each store-week from store-level scanner data (Raju, 1992).

The terms used in the analysis are defined as:

1) *Item* refers to a unique beverage product as identified by Universal Product Code in transaction data. A large supermarket often carries more than 1000 distinct SSB items.

2) *Net price* indicates the price of a SSB item after discounting, standardized to a single serving size (240ml).

3) *Regular price* is the baseline (reference) price of a single serving without any discounting, and

4) *Item* *discounting* indicates the difference between the net and regular price, thus the extent or depth of temporary price reduction per serving. Note that net and regular price are in Canadian Cents.

Because the data provide only the weekly net price after discounting for each item but not the regular price nor discounting directly, we calculated the regular price for each item, and then we computed discounting as the difference between the calculated regular price and the observed net price. We then computed the category-level discounting as the weighted mean of item discounting across items in the same store-week, with the weights representing the market share of each item.

For simplicity, we focus on the calculation of discounting for the soda category as an example. However, the calculations below were applied to each of the five SSB categories. Let $i$ represent an item of soda sold in store $s$ at week $j$. We denote the net price of item $i$ in store $s$ at week $j$ as $P_{ijs}$. We examined the price history of each item. The regular price of soda item $R_{ijs}$ is identified as the highest price in a 3-month window, e.g., a moving maximum net price of item $i$ in store $s$ in a 3 months widow. Supplementary Figure S4-1 below illustrates a series of weekly net price of one of popular beverage items and its regular price.

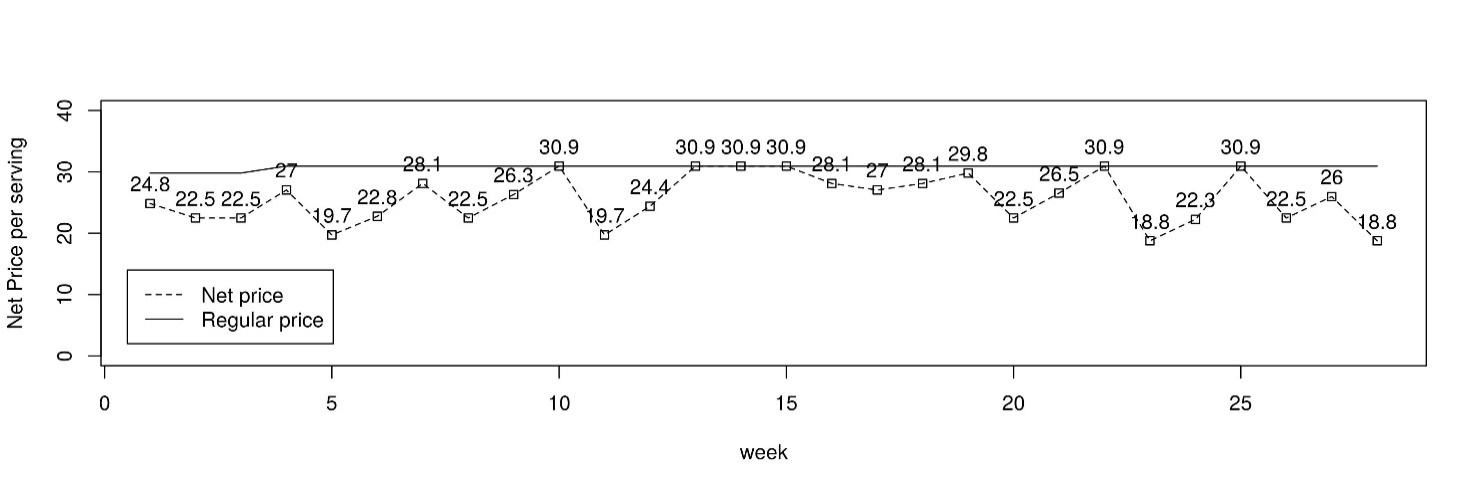

Supplementary Figure S1. Weekly Net Price and Regular Price of a Single Soda Item in Cents.

Item discounting, $D_{ijs}$ , is defined as a percent price decrease from regular price as shown below;

$D_{ijs}$= ${(R}_{ijs}$ – $P_{ijs})$ / $R_{ijs} \times100$

Supplementary Figure S1 indicates that there is no item discounting (zero percent item discounting) in weeks 10, 13-15, 23, and 25, so the net price is the same as the regular price for these weeks. In other weeks, there is discounting since the net price is less than the regular price. In week 11, for example, the discounting is computed as: (30.9 – 19.7) / 30.9 x 100 = 36.25 percent.

Finally, we aggregated the individual item discounting into an overall measure for the discounting in the each of five SSB categories. Suppose there are 300 soda items sold in store $s$ in week $j$. Our main exposure of interest, the category-specific store-week discounting of all soda items, $X_{js}$,is defined as

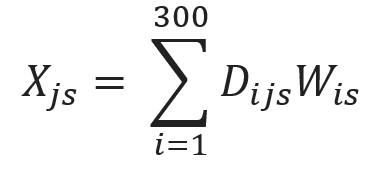
,

where $W_{is}$ represents a normalized (sum to one across all items in store $s$) weight calculated from the annual market share of each soda items within the soda category at store $s$. Therefore, the value of the weights is large and stable over time for soda items of major brands.

### Appendix S3. Shapes of promotional lag

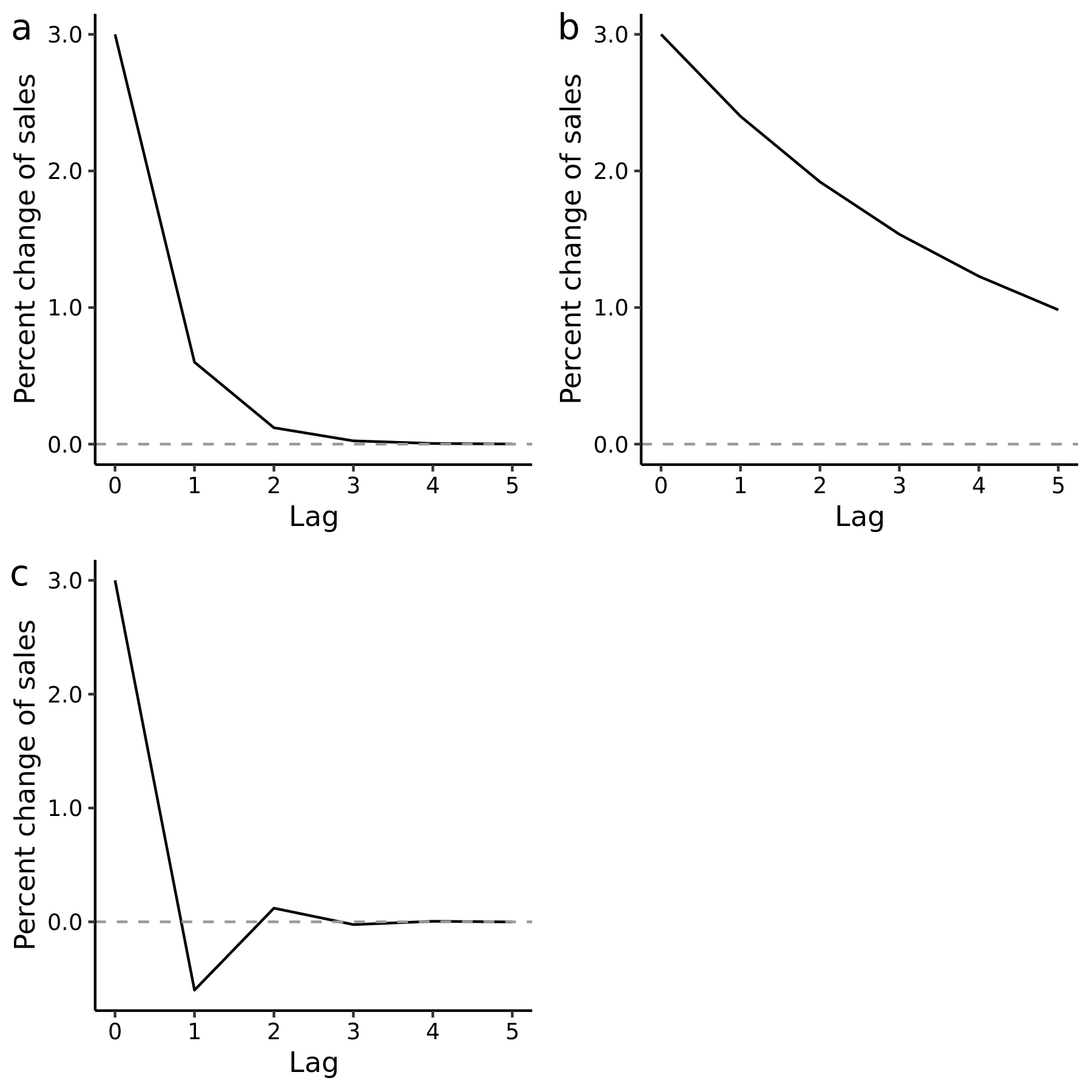

Supplementary Figure S2: Hypothetical shapes of the lagged effect of price discounting as a) a monotonic decay returning to the original (baseline, pre-promotion) level of sales with a weak estimated lag effect ($\lambda=0.2)$, and b) a more persistent lag ($\lambda=0.8)$ such that sales have not decayed to pre-discounting levels after 5 weeks.

The functions seen in plot a) and b) are generated when the value of $\lambda$ is constrained to be between 0 and 1 by an analyst.

Discounting in this plot represents a ‘shock’ exposure, that is, its value is increased by one unit (i.e., one percent discounting) at the week where *x* = 0 (promotion period), then set to zero, or returning to the baseline, pre-discounting, price in the following weeks.

The y-axis represents the effect of one percent discounting on sales at each week. The value at *x* = 0 represents the immediate effect, which is the change of sales during the week of discounting, as represented by the value of $\beta$.

### Appendix S3. Statistical analysis: time-series regression to incorporate the lagged effect of discounting, covariates, and intercept

*Transfer function*

First, we specify the koyck lag specification discussed in the main text. The monotonic decay of discounting at time *t* is summarized by a structural variable $E_{t}$ as:

$E_{t}= \lambda^{0}\beta x_{t} + \lambda^{1}\beta x_{t-1} + \lambda^{2}\beta x_{t-2} +...+ \lambda^{h}\beta x_{t-h}$,

where $x_{t}$ represents price discounting at time *t*, $\beta$ is the coefficient for the immediate effect of discounting, $\lambda$ is the decay parameter as described in the main document, and *h* indicates lag length. The equation above allows the effect of discounting to geometrically decay over lag. Using transfer function, the same equation can be recursively summarized as:

$E_{t}= \lambda E_{t-1} + \beta x_{t}$, which indicates that the structural variable $E_{t}$ captures immediate effect of $x_{t}$ and its cumulative effect up to time *t* through lagged variable $E_{t-1}$ (1). We have provided further details about transfer functions (2).

*Time-series model*

We added the structural variable $E_{t}$to a dynamic liner model, a time-series linear regression model that naturally incorporates time-varying parameters, crucially $E_{t}$ above, to smoothly change over time under structural formulation (1,3). In our analysis, the model is specified as:

$$y_{t}=\alpha_{t}+{Season}_{t}+\gamma_{covariates}C_{t}+E_{t}+\varepsilon$$

$E_{t}= \lambda E_{t-1} + \beta x_{t}$ ,

where $E_{t}$ is the structural variable capturing time-varying influence of discounting combining the immediate and lagged effects up to time *t* as descried in the main text. The outcome is the natural log-transformed sales outcome of each SSB category denoted as $y_{t}$.

The intercept $\alpha_{t}$ is a ‘dynamic’, or time-varying intercept, which captures long-term trends and temporal fluctuations of sales. This is a series of $\alpha$s (one for each week, and thus denoted as $\alpha_{t}$), interpreted as baseline sales at time *t* and is determined by its previous value, $\alpha_{t-1}$and random noise, hence this process is called random walk. Further details in the “local level model” specification are available in references (1,3).

The term $C_{t}$ represents a vector of covariates described in the main document, the corresponding vector of time-fixed coefficients is denoted as $\gamma_{covariates}$. The term ${Season}_{t}$ consists of nonlinear regressors as defined by sine and cosine functions fitted to the harmonic wave, with its regression coefficients controlling amplitude of the seasonal wave. Its periodicity is $2\pi t\omega$, where $t=1,2\ldots, 311$ as the indicator of week, $\omega$ as 52.2 representing the cycle (number of weeks in a year) and $\pi$ = $3.1415\ldots$ as the constant pi. With the regression coefficients, $\gamma_{cos}$ and $\gamma_{sine}$ defining the estimated amplitude (vertical extent).

We used Stan statistical software to fit the models (4). The value of the discounting variable as well as all covariates except dummy indicators for holiday were mean centered to improve the mixing of the Markov chain for successful mixing. We generated 3,000 Markov chains as burn-in sample, followed by 30,000 chains as the posterior distribution for inferencing. Stan codes to the model are available in:

https://github.com/hiroshimamiya/promotionLag/blob/main/discountLag_KoyckTransfer.stan.

*Prior probabilities*

The natural log-transformed sales outcome $Y_{t}$ follows a normal distribution, with its error terms $\varepsilon$ following a zero-mean non-informative (i.e., diffuse) Cauchy distribution $\varepsilon^{+} \sim Cauchy(0, 5^{2})$, which is a commonly suggested distribution for a variance parameter (5). The superscript ^+^ indicates that the distribution is constrained to take a positive value (i.e., constrained to the positive half of the Cauchy distribution). We also used a positive-constrained normal distribution to examine posterior sensitivity with the same values of the scaling parameter as above, which led to nearly identical results. The prior probability of the regression coefficients, including the season coefficients was specified as independent non-informative normal distributions $\beta_{covariates}, \beta_{Cos}, \beta_{Sine}, \beta\sim Normal(0,5^{2})$. For the time-varying intercept, $\alpha_{t}$, the prior probability of the initial value at $t = 1$ was $\alpha_{1}\sim Normal(0,5^{2})$, which was subsequently allowed to evolve as was $\alpha_{t}\sim Normal(\alpha_{t-1},\epsilon_{alpha})$ with a random noise $\epsilon_{alpha} \sim N(0,1)$.

### Appendix S4

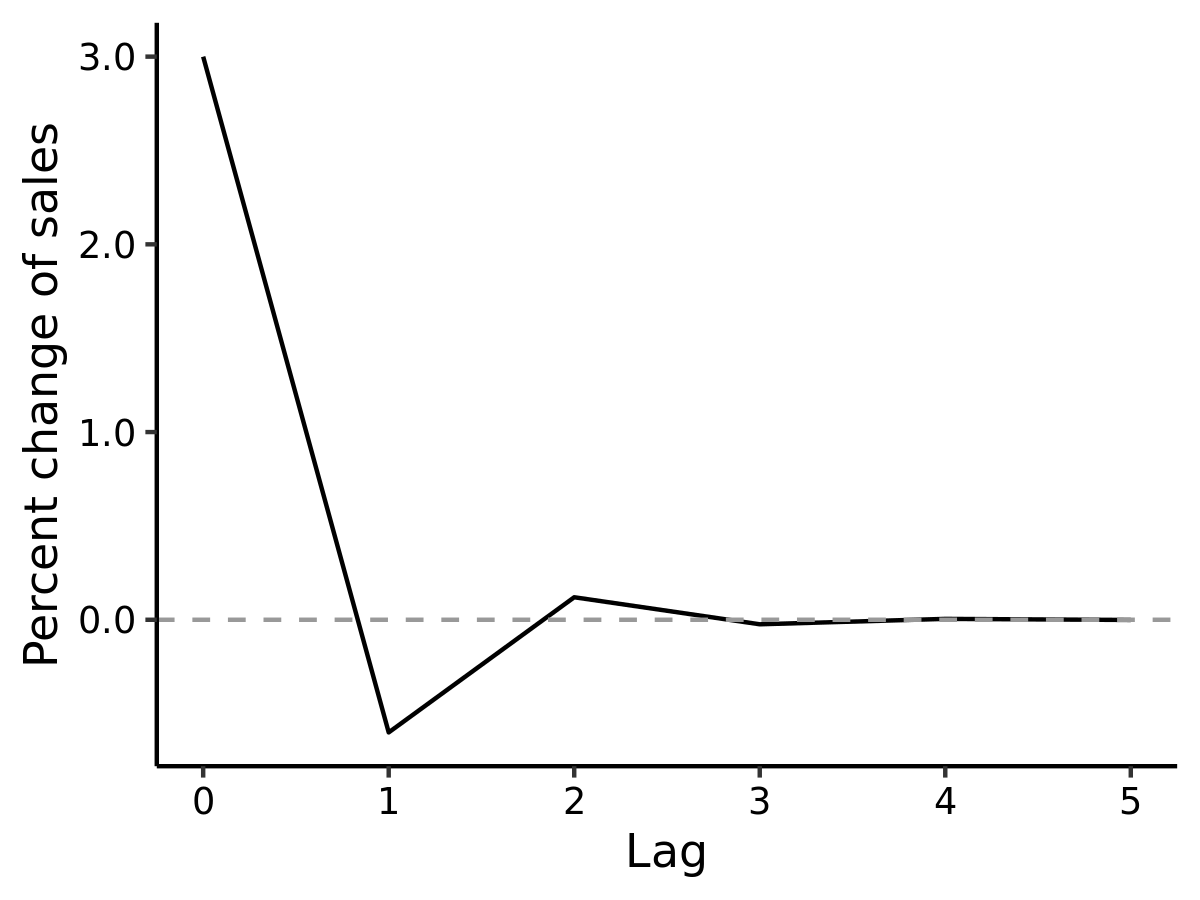

Supplementary Figure S3. Post-promotion dip of sales at one week after the discounting period (*x* = 0).

The y-axis represents the percent change of sales relative to the baseline (pre-discounting) sales due to one percent discounting. Unlike the monotonic decay seen in Supplementary Figure S2 a and b, the oscillating pattern described here is captured by constraining the value of $\lambda$ as: $-1 <\lambda<0$. The decay function in this plot is generated by $\lambda=-0.2.$The dip is attributed to reduced purchasing activities if households stockpile the promoted items during the discounting period. However, the dip may not be observed when stockpiled items are consumed rapidly, leading to an immediate re-purchasing after discounting (6–8).

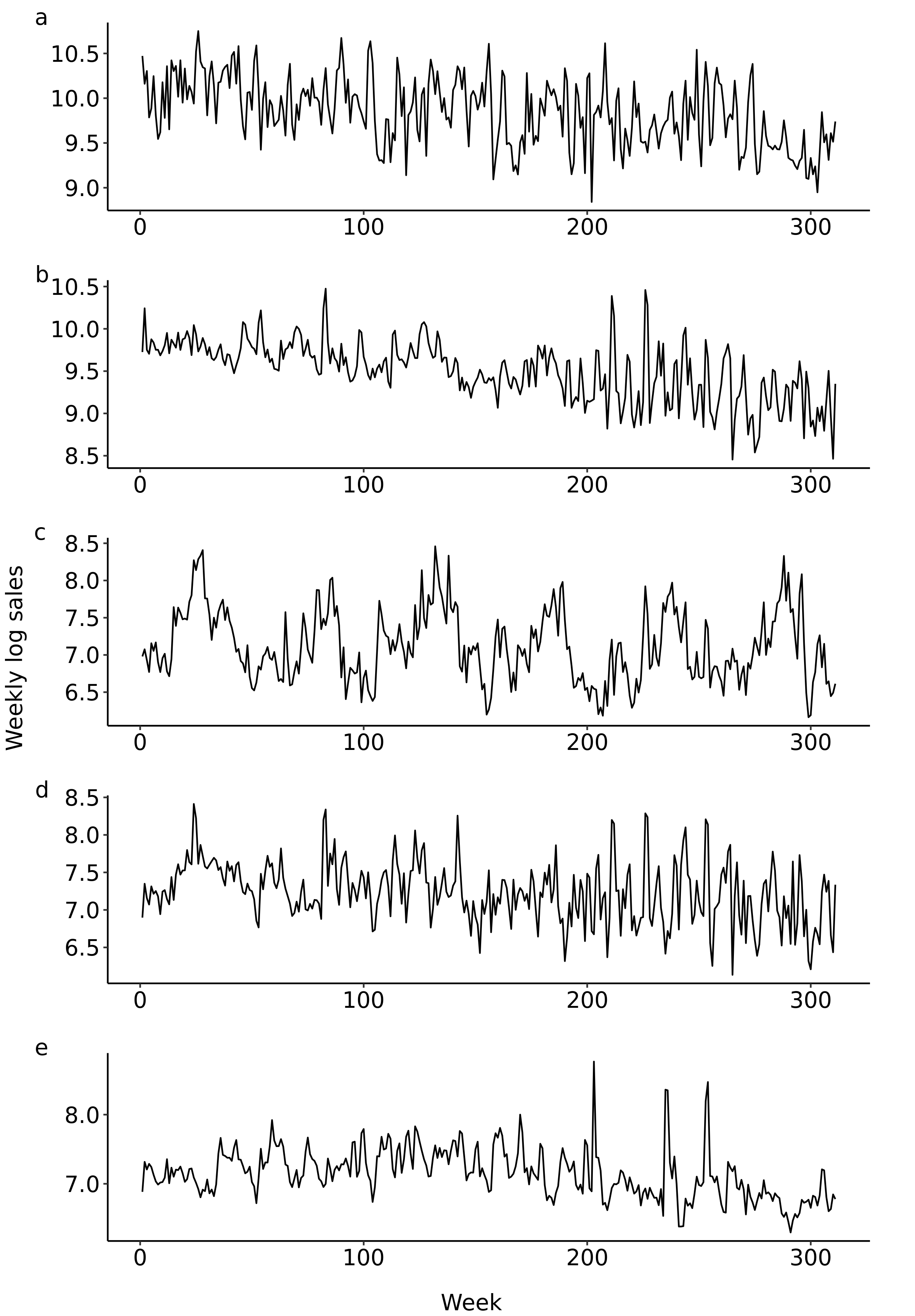

Supplementary Figure S4: Weekly series of category-level percent sales for a) soda, b) fruit drinks, c) sports and energy drinks, d) coffees and teas, and e) drinkable yogurt.

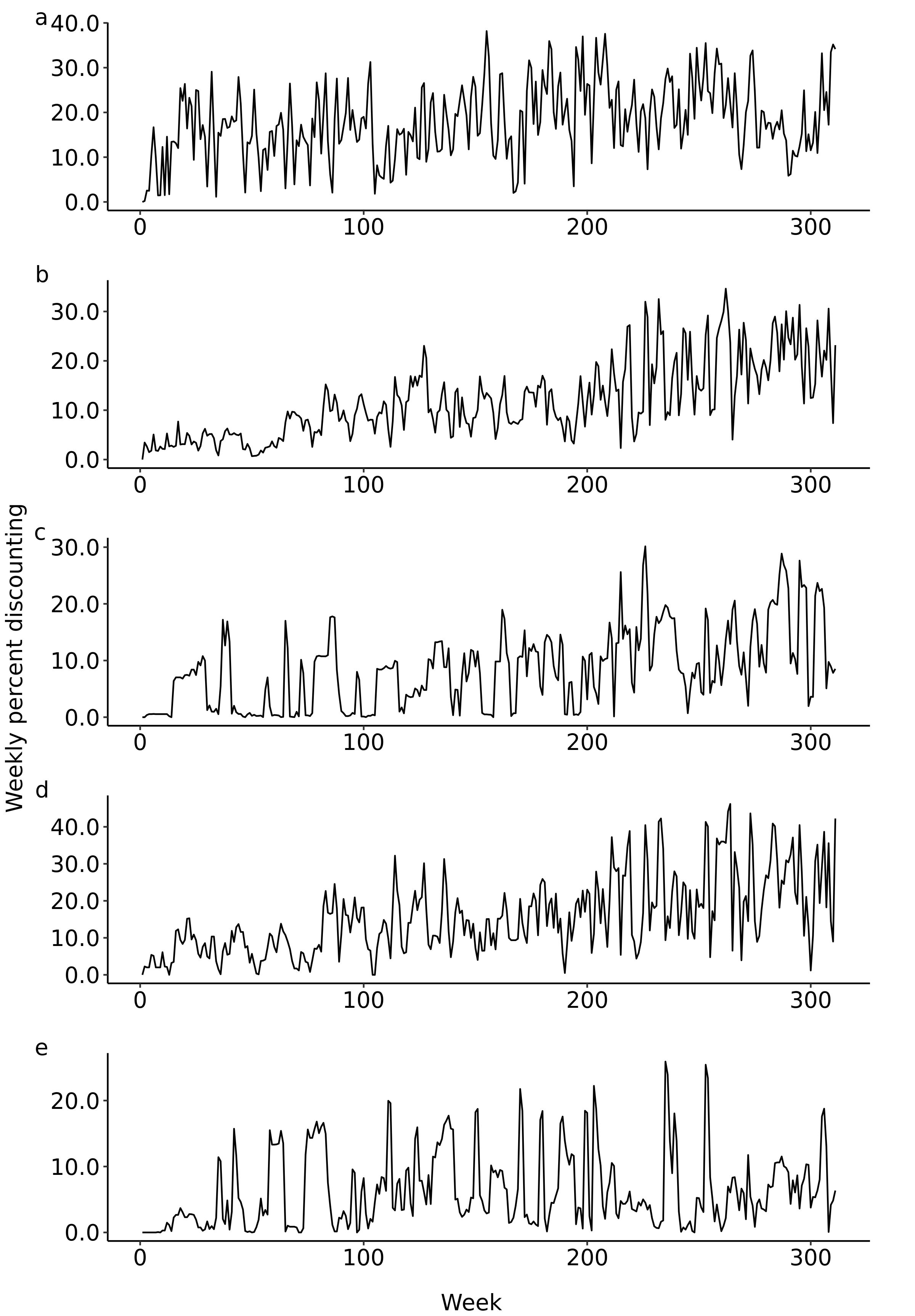

Supplementary Figure S5: Weekly series of category-level percent discounting for a) soda, b) fruit drinks, c) sports and energy drinks, d) coffees and teas, and e) drinkable yogurt.

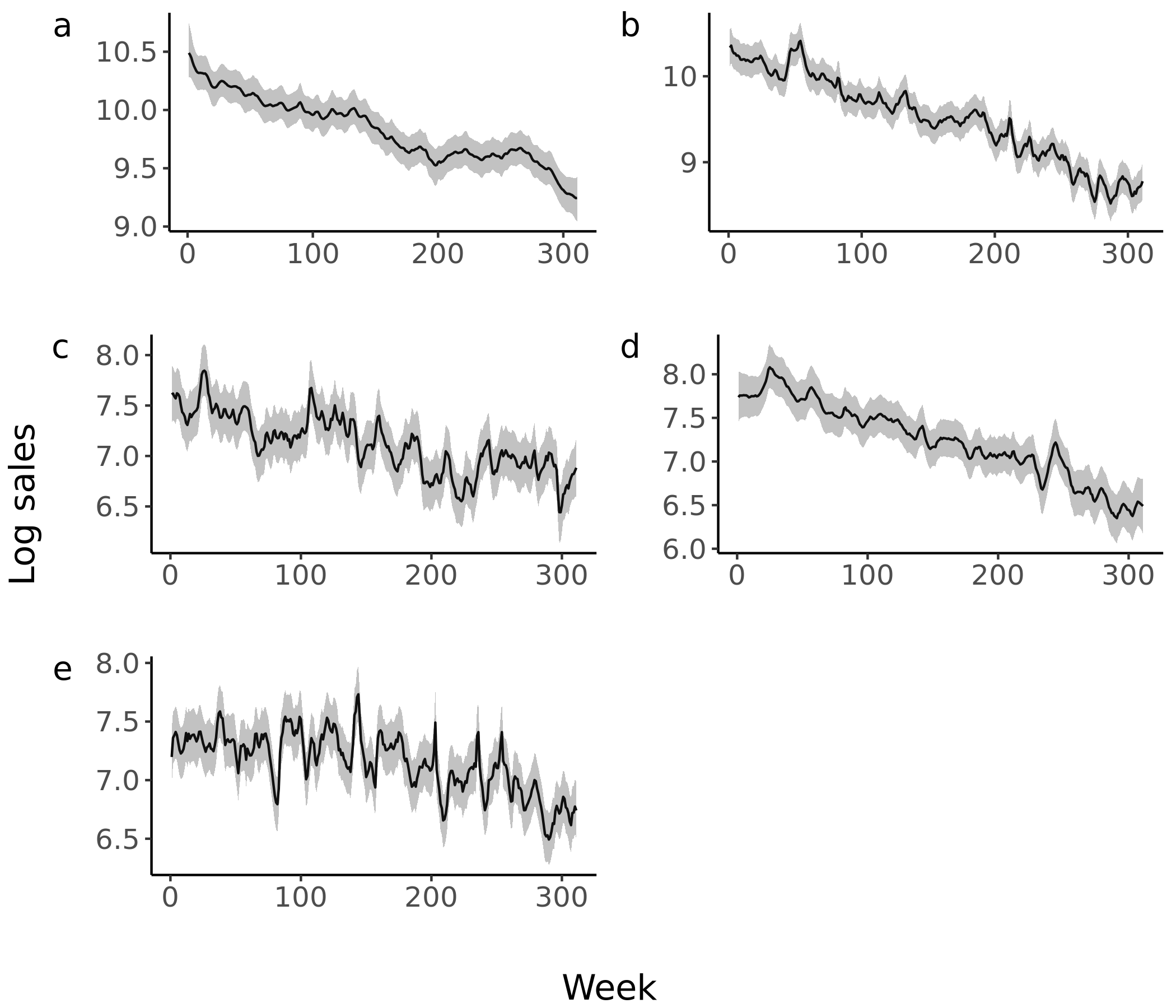

Supplementary Figure S6. Posterior mean (solid line) and 95% credible interval (grey band) of weekly-varying intercept, $\alpha_{t}$, of log sales for a) soda, b) fruit drinks, c) sports and energy drinks, d) coffees and teas, and e) drinkable yogurts. Note that the value of the discounting variable was mean centered; therefore, the estimated level of the intercept represents the sales at the mean level of discounting.

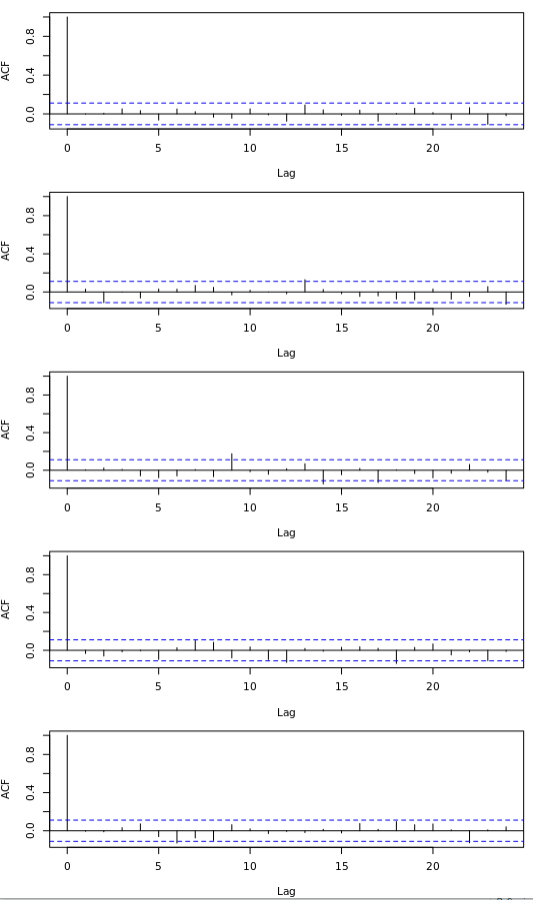

Supplementary Figure S7**.** Autocorrelation function of residuals for : a) soda, b) fruit drinks, c) sports and energy drinks, d) coffees and teas, and e) drinkable yogurt category. The grey band around solid line indicates 95% credible interval. It should be noted that the fitted sales in these plots do not represent forecasting distribution.

Supplementary Table 2. Non-log serving-standardized sales quantity of SSB categories attributed to lagged effect along (left pane) and both lagged and immediate effect (right pane).

|  | Sales due to lagged effect | | |  | Sales due to lagged and immediate effect | | |
| --- | --- | --- | --- | --- | --- | --- | --- |
| SSB category | Mean | Median | 95%CI |  | Mean | Median | 95%CI |
| Soda | 306,483 | 238,042 | (9,943- 977,152) |  | 4,308,369 | 4,283,077 | (3,418,117 - 5,343,486) |
| Fruits drinks | 195,659 | 174,491 | (10,659 - 508,513) |  | 2,317,649 | 2,306,233 | (1,876,257 - 2,824,259) |
| Energy and sports drinks | 28,459 | 26,345 | (2,661 - 67,253) |  | 131,606 | 130,446 | (96,155 - 173,625) |
| Sweetened coffees and teas | 20,079 | 16,627 | (802 - 58,650 |  | 281,883 | 279,539 | (216,303 - 360,958) |
| Sweetened drinkable yogurts | 6,007 | 4,870 | (213- 18,060 |  | 133,117 | 132,531 | (108,460 - 161,073) |
